# Assessment of Health-Related Quality of Life (HRQoL) of Type 2 Diabetic Patients in Ghana

**DOI:** 10.1101/2024.09.28.24314542

**Authors:** Christine Ahiale, Augustine Kumah

**Author notes:** **Corresponding Author:** Augustine Kumah, Research on Interventions for Global Health Transformation-RIGHT Institute.

## Abstract

**Background:** Over the last decades, non-communicable diseases such as diabetes have remarkably increased due to rapid urbanization, unhealthy lifestyles, and ageing. The World Health Organization (WHO) indicates that the “diabetes epidemic” will continue in the coming decades, yielding enormous human and economic costs around the world. This study aims at accessing the health-related Quality of Life (QoL) of Type 2 Diabetic Patients in Ghana.

**Method:** This was a hospital-based cross-sectional descriptive study conducted in the 3 regions in Ghana among 1194 patients with type 2 diabetes who were seeking healthcare at the regional hospitals of the selected regions for at least 12 months. A multi-stage sampling technique was used in recruiting the participants. The WHOQOL-BREF tool was adopted to assess the health-related QoL of diabetic patients. Data was analyzed using STATA version 16.1, and the findings were presented in the tables. Statistical significance was set at (P < 0.05).

**Results:** Out of the 1194 participants, 90% (1079) reported a satisfactory quality of life. The majority (60.3%) of participants were above 50 years of age. The overall mean score of the QoL was 6.25 (±1.28) for all four domains: psychological, physical, social, relationship, and environmental health. Marital status, occupational status, and self-reported health status predicted overall QOL. Males have a better quality of life than females, and those below 50 also have a better quality of life than those above 50. Similarly, those employed had a better quality of life than the unemployed; however, those who were married, unmarried, educated, and non-educated had no significant differences in their quality of life. Sex was associated with all the domains. Marital status was associated with the environmental, social, and psychological domains. Age was associated with psychological and physical domains. On predictors of quality of life, all demographic variables (age, sex, educational status, marital status, and employment status) predicted quality of life across all domains.

**Conclusion:** This study provided valuable insights on predictors of health-related QoL among type 2 diabetic patients in Ghana, which would guide policymakers, healthcare providers, and stakeholders in improving diabetes management and ultimately enhancing the quality of life for individuals living with diabetes in Ghana. Evidence-based interventions should be implemented to address the factors influencing the QoL of diabetic patients in Ghana.

## Introduction

Diabetes Mellitus (DM) is characterized by elevated blood sugar levels and is classified as a chronic metabolic disorder according to the World Health Organization^1^. Normal fasting blood sugar levels are 99 mol/dL or lower. In comparison, levels between 100 to 125 mg/dL indicate prediabetes and 126 mg/dL or higher indicate the presence of DM, as per the Centers for Disease Control and Prevention^2^. Failure to seek timely medical interventions can lead to complications.

The global prevalence of type 2 DM is projected to increase to 7079 individuals per 100,000 by 2030, reflecting a continued rise across all regions of the world^3,4^. The WHO ranked DM as the 9^th^ leading cause of death globally^1^. Moreover, the hospitalization and mortality rates from type 2 DM are increasing across countries of all economic income. In Nigeria, for instance, the hospital admission rate for type 2 DM was estimated at 222.6 per 100,000 population with hyperglycemic emergencies^5^. The overall mortality rate was 30.2 per 100,000 population, with a case fatality rate of 22.0%^5^.

In Ghana, numerous studies conducted on the prevalence of Type 2 Diabetes Mellitus (T2DM) in various geographical locations have consistently indicated a high prevalence^6–8^. A study in the Central Belt of Ghana revealed a substantial increase in type 2 diabetes admission rates, rising from 2.36 per 1000 admissions in 1993 to 14.94 per 1000 admissions in 2014, representing a staggering 633% surge over the 31-year period^9^. Over the same 31-year period, in-patient diabetes fatality rates witnessed an increase from 7.6 per 1000 deaths to 30 per 1000 deaths^9^ additionally, a retrospective cross-sectional study conducted on predominant complications of type 2 DM in Ghana revealed that the prevalence of macrovascular and microvascular complications of type 2 DM was 31.8% and 35.3% respectively^10^. The study further revealed that the prevalence of neuropathy, nephropathy, retinopathy, sexual dysfunction, diabetic ketoacidosis, and hypoglycemia were 20.8%, 12.5%, 6.5%, 3.8%, 2.0%, and 0.8% respectively^10^. All these complications of type 2 DM can be minimized if much effort is directed toward improving the QoL of people with type 2 DM.

Due to the impossibility of complete treatment of chronic diseases, assessing the quality of life (QOL) among patients with such diseases is a vital outcome measure^11^. According to the definition proposed by WHO, QOL is defined as “individuals’ perception of their position in life in the context of the culture and value systems in which they live and concerning their goals, expectations, standards, and concerns^12^. In most health-related literature, the term “health-related quality of life” (HRQOL) is gradually accepted instead of QOL, a multi-dimensional structure of subjective evaluation of the good life, including performance concerning physical, mental, and social subjects.

QOL among people with diabetes has been assessed in some studies conducted in different regions using different instruments and methodologies. Various standardized tools, such as the Diabetes Quality of Life Measure (DQOL) and the EuroQol-5D (EQ-5D), are used to assess HRQoL in T2DM patients. These instruments provide valuable insights into the physical, mental, and social dimensions of QoL and guide interventions to improve patient well-being^11^. A study among diabetic patients in Western Ethiopia revealed a mean score of 50.3+-18.1 for overall HRQoL^13^. There was a poor HRQoL (30.2+-22.9) in the general health domain and a high HRQoL (63.2+- 34.4) in the physical functioning area^13^. Among individuals with diabetes, public health, mental health, physical discomfort, and vitality are the HRQoL domains most negatively impacted, with a mean score of less than 50^13^. A higher QoL score of 74.1+11.6 was discovered in another cross-sectional study on the determinants of QoL and glycemic control among Saudi individuals with diabetes^14^. Additionally, a cross-sectional study involving individuals with QoL predictors and other patient-reported outcomes of type 2 diabetes indicates a good QoL of a score of 0.93, which approximates good^15^.

A unit increase in age is likely to decrease the HRQoL of a patient with diabetes by 0.25, according to a cross-sectional study on predictors of HRQoL among patients with diabetes on follow-up in Western Ethiopia^13^. Compared to female diabetes patients, male patients had an HRQoL roughly five times higher^13^. Married diabetes patients had a five-fold higher HRQoL than single patients, and people without literacy skills had a nine-fold lower HRQoL (3.6 units) than people with literacy skills^13^. Those with a history of smoking had HRQoL that was nine units lower than those without a history of smoking, even in the absence of comorbidity and Diabetes-related chronic complications raise HRQoL, and HRQoL is expected to drop by 3.56 for every unit increase in body mass index (BMI)^13^. Another cross-sectional study on QoL predictors in diabetes mellitus patients at two tertiary health institutions in Ghana and Nigeria reveals that medication adherence, employment status, and diabetes empowerment are the main factors^16^. Higher medication adherence was linked to an improvement in type 2 diabetes patients’ HRQoL, according to a cross-sectional descriptive study on the profile and predictors of HRQoL among patients with type 2 diabetes mellitus in Pakistan^17^. There is a significant correlation between age, treatment satisfaction, medication adherence, BMI, and HbA1c and HRQoL predictors for type 2 diabetes patients using insulin, according to another cross-sectional observational study on the subject^18^. This study, therefore, seeks to assess the health-related Quality of Life (QoL) of Type 2 Diabetic Patients in Ghana.

## Methods

### Study design

This study utilized a facility-based cross-sectional descriptive research design to assess the health-related quality of life (HRQoL) of Type 2 Diabetic (T2DM) patients in Ghana between January and May 2023.

### Study Area

This study was carried out in 3 regions across the three ecological belts in Ghana: Northern Region, Ashanti Region, and Greater Accra Region. These regions were purposefully selected because they are the most populated regions in the northern, middle, and southern belts, respectively, and have the highest burden of diabetes in Ghana.

### Study Population

All persons aged 18 years and above who were T2DM, had been medically diagnosed for at least one year, and were receiving care at the diabetic clinics of the three selected regional hospitals of the respective regions at the time of this study were included.

### Sampling method

This study used a multi-stage sampling method. First, three regions and their regional hospitals were purposefully selected. The diabetic clinic registers in each selected facility were obtained and screened to identify patients diagnosed with T2DM who met the inclusion criteria. A simple random sampling technique was used to sample participants proportionate to the size of the selected health facilities. In all, 1194 T2DM patients were recruited for the study.

#### Study Material

The WHOQOL-BREF tool was adapted to assess the QoL of people with T2DM. WHOQOL-BREF is a shortened version of the WHOQOL-100 tool^19^. The WHOQOL-BREF instrument comprises four primary domains: physical, psychological, social, and environmental. Besides these four domains, two items were examined separately: an individual’s overall perception of QoL and overall perception of his or her health^19^.

### Data analysis

The data was collected using a Kobo Collect, exported as an Excel file, and analyzed using STATA version 20.1. Regarding the WHOQOL-BREF data, where up to two items are missing, the mean of other items in the same domain will be substituted^12^. Also, where more than two items are missing from the domain, the domain score will not be calculated (except for domain 3, where the domain should only be estimated if up to one item is missing)^12^.

To assess QoL, 4 domain scores that measure overall QoL were computed. The scores in the four domains (physical, psychological, social, and environmental) represent an individual’s quality of life (QoL) assessment. These scores are oriented positively, meaning higher scores indicate a higher QoL. The mean score of the items within each domain was determined to calculate the domain score. These mean scores were multiplied by 4 and subsequently transformed to a 10-100 scale using a specific formula recommended by the WHO (2015).

In the study context, higher scores on the quality of life (QoL) assessment indicated a better QoL for the participants. The socio-demographic characteristics of the diabetic patients were presented in the form of percentages, frequencies, and means, which helped provide a comprehensive overview of the participant population. QoL scores were presented as mean ± SD. Pearson’s correlation coefficient was used to assess the inter-domain correlation and the correlation between various demographic factors and domain scores. Multiple regression analysis was done to determine independent predictors of QoL. A p-value of ≤ 0.05 was considered statistically significant. Socio-demographic data was used as predictor variables, and all four domains, including items 1 and 2, were used as outcome variables.

### Ethical Considerations

Ethical clearance was obtained from the Ghana Health Service Ethics Review Committee and the University of Port Harcourt Research Ethics Review Board. Before the commencement of the study, permission was obtained from the regional health directorates and hospital administrators of all selected regions and hospitals, respectively. Informed consent was obtained from each participant before the interviewer, and the participants were allowed to withdraw from the study at any time without penalty.

## Results

### Socio-Demographic Characteristics

In all, 1194 participants were involved in this study. With a mean age and standard deviation of 54.23 (±13.41), the study revealed that the majority (60.3%) of participants were above 50 years of age. About 5% were less than 30 years of age. For sex, 56.5% of study participants were males. In terms of education, more than a quarter (25.5%) of participants had a tertiary level of education. Also, more than half (61.9%) of participants were married during the study. More than half (50.1%) of study participants were employed. Out of this, 62.5% were working in the private sector. Most (41.6%) employed participants received less than 500.00 Cedis monthly (Table 1).

**Table 1:** Socio-Demographic Characteristics of Type II Diabetes Patients.

| Variables | Frequency (N=1194) | Percentage (%) |
| --- | --- | --- |
| **Age Group** |  |  |
| • < 30 years | 71 | 5.9 |
| • 30 - 50 years | 403 | 33.8 |
| • > 50 years | 720 | 60.3 |
| **Mean Age (±SD)** | **54.23 ($\pm 13.41$)** |  |
| **Sex** |  |  |
| • Male | 675 | 56.5 |
| • Female | 519 | 43.5 |
| **Educational status** |  |  |
| • None at all | 222 | 18.6 |
| • Primary school | 106 | 8.9 |
| • Junior High School | 319 | 26.7 |
| • Senior High School | 243 | 20.4 |
| • Tertiary | 304 | 25.5 |
| **Marital Status** |  |  |
| • Living as married | 739 | 61.9 |
| • Divorced | 49 | 4.1 |
| • Separated | 104 | 8.7 |
| • Single (never married) | 144 | 12.1 |
| • Widowed | 158 | 13.2 |
| **Employment status** |  |  |
| • Employed | 598 | 50.1 |
| • Unemployed | 596 | 49.9 |
| **Type of employment** |  |  |
| • Government worker | 114 | 19.1 |
| • Private sector worker | 110 | 18.4 |
| • Private sector worker | 374 | 62.5 |
| **Estimated monthly income** |  |  |
| • <500 | 249 | 41.6 |
| • 500 – 1000 | 197 | 32.9 |
| • 1100 – 2000 | 87 | 14.6 |
| • > 2000 | 65 | 10.9 |
| **Median (IQR) income** |  | **600 (400 – 1150)** |
| **Health status** |  |  |
| • Healthy | 583 | 48.8 |
| • Unhealthy | 611 | 51.2 |

IQR (Interquartile range), SD (Standard Deviation)

### Assessing the health-related quality of life of T2DM Patients in Ghana

When evaluating the participants’ health-related quality of life, it was found that out of the total 1194 individuals, 90% (1079) reported a satisfactory to good quality of life. The remaining 10% (115) indicated poor health-related quality of life. (Table 1 and Table 2)

**Table 2:** Quality of Life per Age and Gender.

| Quality of life | Age ≤ 50 |  | Age > 50 |  | Female |  | Male |  |
| --- | --- | --- | --- | --- | --- | --- | --- | --- |
|  | No | % | No | % | No | % | No | % |
| Poor | 23 | 5.8 | 92 | 11.5 | 31 | 6.0 | 84 | 12.4 |
| Satisfactory | 193 | 48.7 | 416 | 52.1 | 281 | 54.1 | 328 | 48.6 |
| Good | 180 | 45.5 | 290 | 36.4 | 207 | 39.9 | 263 | 39.0 |
| Total | 396 | 100 | 798 | 100 | 519 | 100 | 675 | 100 |

Among those reporting satisfactory health-related quality of life, males constituted the majority at 55% (591). Regionally, good quality of life relating to health was found among the Ashanti Region (47%), followed by greater Accra (33%) and the Northern Region (32%). Out of 115 with stated poor health-related quality of life were among those in the capital city of Greater Accra region, scoring (56%), 38%, and 6% by Northern and Ashanti Regions, respectively. (Table 3)

**Table 3:**
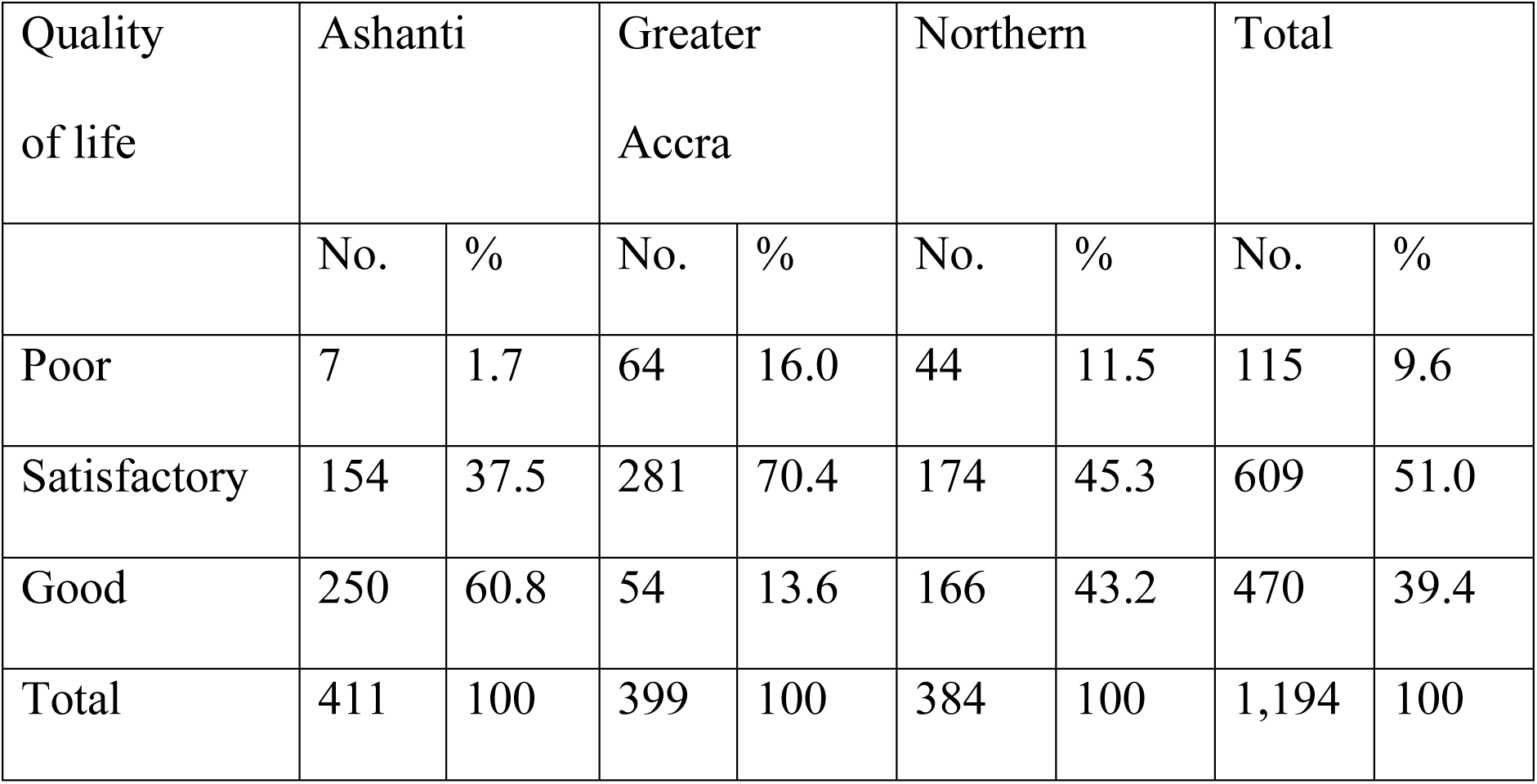
Quality of life by region.

| Quality of life | Ashanti |  | Greater Accra |  | Northern |  | Total |  |
| --- | --- | --- | --- | --- | --- | --- | --- | --- |
|  | No. | % | No. | % | No. | % | No. | % |
| Poor | 7 | 1.7 | 64 | 16.0 | 44 | 11.5 | 115 | 9.6 |
| Satisfactory | 154 | 37.5 | 281 | 70.4 | 174 | 45.3 | 609 | 51.0 |
| Good | 250 | 60.8 | 54 | 13.6 | 166 | 43.2 | 470 | 39.4 |
| Total | 411 | 100 | 399 | 100 | 384 | 100 | 1,194 | 100 |

### Comparison of the WHOQOL-BREF mean scores in four domains according to sex, age, education status, marital status, employment status, and health status

The study conducted a comparison of the WHOQOL-BREF mean scores in four domains according to sex, age, education status, marital status, employment status, and health status, and this is presented in the table below. From Table 4, there was an increased mean score for all four domains (physical, psychological, social, and environmental) of health and the overall quality of health among participants less than 50 years compared to participants 50 years and beyond. Similarly, males had higher mean scores for all four domains and the overall quality of life than females. For instance, males had a mean score of 22.52 and SD of 3.16 compared to the mean score and Standard deviation (SD) of 21.29±2.98 for females. Also, educated participants had higher mean scores compared to participants who were not educated. Married participants had a higher mean score in all domains than unmarried/divorced/widowed participants. In addition, employed participants had higher mean scores for all four domains than unemployed participants. (Table 4)

**Table 4:** Comparison of the WHOQOL-BREF mean scores in four domains according to sex, age, education status, marital status, employment status, and health status.

|  | Domains |  |  |  |  |
| --- | --- | --- | --- | --- | --- |
| | Physical health<br>Mean ±SD | Psychological<br>health<br>Mean ±SD | Social<br>relationships<br>Mean ±SD | Environmental<br>Health<br>Mean ±SD | Overall QoL<br>Mean ±SD |
| **Age Group** |  |  |  |  |  |
| <50 year | 23.21±2.97 | 19.66±2.78 | 10.01±1.68 | 26.00±3.44 | 6.49±1.19 |
| ≥50 year | 20.91±2.88 | 18.08±2.95 | 9.56±1.91 | 25.32± 3.43 | 6.08± 1.31 |
| P-value | <0.001 | <0.001 | <0.001 | 0.0008 | <0.001 |
| **Sex** |  |  |  |  |  |
| Female | 21.29±2.98 | 18.26±2.94 | 9.59± 1.91 | 25.17± 3.49 | 6.11± 1.35 |
| Male | 22.52±3.16 | 19.29±2.95 | 9.93± 1.71 | 26.14±3.32 | 6.42± 1.15 |
| P-value | 0.001 | 0.0000 | 0.0012 | <0.001 | 0.001 |
| **Educational level** |  |  |  |  |  |
| Not educated | 21.62±3.23 | 18.96±3.18 | 9.63±1.79 | 25.25± 3.56 | 6.06± 5.89 |
| Educated | 21.87±3.09 | 18.65± 2.94 | 9.76±1.99 | 25.67±3.42 | 6.29± 6.21 |
| P-value | 0.2866 | 0.1828 | 0.3973 | 0.1066 | 0.0156 |
| **Marital Status** |  |  |  |  |  |
| Married | 21.88±2.89 | 18.96±2.91 | 9.99±1.77 | 25.94±3.32 | 6.37± 1.25 |
| Single/Divorced/Widowed | 21.73±3.35 | 18.29±3.05 | 9.32±1.86 | 25.03±3.58 | 6.05± 1.29 |
| p-value | 0.4206 | 0.010 | <0.001 | <0.001 | 0.004 |
| **Employment status** |  |  |  |  |  |
| Employed | 22.88±2.65 | 19.75±2.49 | 10.26±1.66 | 26.35±3.28 | 6.59± 1.23 |
| Unemployed | 20.76±3.19 | 17.66±3.07 | 9.21±1.85 | 24.83±3.45 | 5.83±2.06 |
| P-value | <0.001 | <0.001 | <0.001 | 0.003 | <0.001 |
| **Health status** |  |  |  |  |  |
| Ill health | 22.38±3.33 | 19.23±2.86 | 9.73± 1.73 | 25.61±3.52 | 6.43± 1.15 |
| Healthy | 21.29±2.81 | 18.21±3.02 | 9.74±1.93 | 25.57±3.38 | 6.06± 1.36 |
| P-value | <0.001 | <0.001 | 0.9067 | 0.8431 | <0.001 |

The study found that more females recorded satisfactory and good quality of life (50%) compared to males (41%). Those in the Ashanti Region have a better quality of life than Greater Accra and the Northern Region. (Table 4)

### Predictors of health-related QoL among type 2 diabetic patients. (Table 5)

#### Physical domain

There was a statistically significant association between age, sex, educational status, marital status, employment status, and health status among the study participants. A unit increase in age decreases (Crude β-coef = -0.25, 95% CI: 0.28, -0.21) the probability of having good physical health.

**Table 5:** Predictors of Quality of life.

| | Crude β-coef [95% CI] | p-value | Adjusted β-coef (95% CI) | p-value | R<sup>2</sup> |
| --- | --- | --- | --- | --- | --- |
| **Physical Domain** |  |  |  |  |  |
| • Age | -0.25[-0.28, -0.21] | <0.001 | -0.03[-0.06, -0.01] | 0.004 |  |
| • Sex | 1.07[0.54, 1.40] | <0.001 | - | - |  |
| • Educational status | 0.88[0.36, 1.40] | 0.001 | - | - |  |
| • Marital Status | 1.69[1.18, 2.21] | <0.001 | - | - | 0.25 |
| • Employment status | 3.11[2.59, 3.64] | <0.001 | 1.01[0.63, 1.38] | <0.001 |  |
| • Health status | -5.38[-5.85, -4.91] | <0.001 | -3.08[-3.45, -2.72] | <0.001 |  |
| **Psychiatric Domain** |  |  |  |  |  |
| • Age | -0.12 [-0.15, -0.09] | <0.001 | -0.04[-0.06, -0.01] | 0.004 |  |
| • Sex | 0.16[-0.22, 0.55] | 0.411 | - | - |  |
| • Educational status | 0.39[0.01, 0.77] | 0.042 | - | - |  |
| • Marital Status | 0.49[0.12, 0.87] | 0.010 | - | - | 0.25 |
| • Employment status | 1.62[1.23, 2.01] | <0.001 | 1.01[0.63, 1.38] | <0.001 |  |
| • Health status | -3.39[-3.75, -3.03] | <0.001 | -3.08[-3.45, -2.72] | <0.001 |  |
| **Social Domain** |  |  |  |  |  |
| • Age | -0.09[-0.11, -0.78] | <0.001 | -0.05[-0.06, -0.03] | <0.001 |  |
| • Sex | 0.23[-0.03, 0.48] | 0.081 | - |  |  |
| • Educational status | 0.71[0.46, 0.96] | <0.001 | 0.54[0.31, 0.77] | 0.855 | 0.19 |
| • Marital Status | 0.74[0.49, 0.98] | <0.001 | -0.33[-0.47, -0.19] | <0.001 |  |
| • Employment status | 0.44[1.18, 1.69] | <0.001 | 1.02[0.76, 1.27] | <0.001 |  |
| • Health status | -1.41[-1.66, -1.15] | <0.001 | -1.03[-1.29, -0.78] | <0.001 |  |
| **Environment Domain** |  |  |  |  |  |
| • Age | -0.13[-0.17, -0.09] | <0.001 | -0.07[-0.11, -0.04] | <0.001 |  |
| • Sex | 1.18[0.68, 1.67] | <0.001 | 0.99[0.54, 1.45] | <0.001 |  |
| • Educational status | 0.90[0.41, 1.39] | <0.001 | 0.65[0.20, 1.10] | <0.001 |  |
| • Marital Status | 1.30[0.82, 1.79] | <0.001 | - | - | 0.17 |
| • Employment status | 0.78[0.26, 1.31] | 0.003 | - | - |  |
| • Health status | -3.52[-4.01, -3.03] | <0.001 | -3.07[-3.57, -2.56] | <0.001 |  |
| **General QOL** |  |  |  |  |  |
| • Age | -0.02[-0.02, -0.01] | <0.001 | -0.04[-0.05, -0.02] | <0.001 |  |
| • Sex | 0.30[0.16, 0.45] | <0.001 | 0.27[0.08, 0.46] | 0.005 |  |
| • Educational status | 0.22[0.04, 0.41] | 0.018 | - | - |  |
| • Marital Status | 0.31[0.17, 0.46] | <0.001 | - | - | 0.31 |
| • Employment status | 0.70[0.56, 0.84] | <0.001 | 0.28[0.07, 0.49] | 0.009 |  |

Females were more likely (Crude β-coef = 1.07 95% CI:0.54, 1.40) to have increased quality of life in the physical domain compared to their male counterparts. Also, being married increased the probability of having increased (Crude β-coef = 1.69 95% CI: 1.18, 2.21) quality of life in the physical domain compared to unmarried participants. Employed participants had increased (Crude β-coef =3.11 95% CI: 2.59, 3.64) quality of life in the physical domain compared to unemployed participants. (Table 5)

After adjusting for all other covariates, being employed increased the probability of (Adjusted β-coef =1.0195% CI: 0.63, 1.38) of having improved quality of life in the physical domain compared to being unemployed. Also, a unit increase in age decreases the probability (Adjusted β-coef =- 0.03 95% CI: -0.06, -0.01) of quality of life in the physical domain after adjusting for all other variables. (Table 5)

#### Psychological Domain

A unit increase in age decreases (Crude β-coef= -0.12 95% CI: -0.15 -0.09) psychological health of study participants. Similarly, being married increased (Crude β-coef = 1.69 95% CI: 1.18, 2.21) the quality of life in the psychological domain compared to not being married. Being employed increased (Crude β-coef =1.62 95% CI: 1.23, 2.01) the psychological health of participants compared to unemployed participants. However, participants indicated that they had a reduced chance of increased psychological health (Crude β-coef =-3.39 95% CI: -3.75, -3.03). (Table 5)

The adjusted model revealed that a unit increase in age decreased (Adjusted β-coef =-0.03 95% CI: -0.06, -0.01) psychological health of participants after controlling for all other variables. Similarly, individuals who reported being healthy had reduced (Crude β-coef = -3.08[-3.45, -2.72) psychological health. However, being employed increased (Adjusted β-coef =1.01 95% CI: 0.63, 1.38) the psychological health of participants compared to being unemployed. (Table 5)

#### Social Domain

Education was found to positively impact social domain quality of life, with an increase in quality of life for those who were educated compared to those who were not (Crude β-coef = 0.71, 95% CI: 0.46, 0.96). Similarly, being married was associated with increased social domain quality of life (Crude β-coef = 0.74, 95% CI: 0.49, 0.98) compared to unmarried participants. (Table 5)

Employment status also played a role, as employed participants were more likely to have a higher quality of life in the social domain than unemployed participants (Crude β-coef = 0.44, 95% CI: 1.18, 1.69). On the other hand, there was a negative association between age and social domain quality of life, with a unit increase in age leading to a decrease in quality of life (Crude β-coef = - 0.09, 95% CI: -0.11, -0.78). (Table 5)

After controlling for all other variables, a unit increase in age decreased (Adjusted β-coef = -0.05 95% CI: -0.06, -0.03) the total life quality in the social domain. Similarly, being married decreased (Adjusted β-coef =-0.33 95% CI: -0.47, -0.19) quality of life in the social domain compared to unmarried participants after adjusting for all other variables. However, being employed increased (Adjusted β-coef =1.02 95% CI: 0.76, 1.27) the quality of life in the social domain compared to being unemployed after controlling for all covariates. (Table 5)

#### Environmental Domain

Females were more likely to have increased (Crude β-coef = 1.18 95% CI: 0.68, 1.67) better life well-being in the environmental domain compared to males. Being employed increased the likelihood (Crude β-coef =0.78 95% CI: 0.26, 1.31) of having a high quality of life in the environmental domain compared to unemployed participants. However, a unit increase in age decreased (Crude β-coef = -0.13 95% CI: -0.17, -0.09) the probability of having a high quality of life in the environmental domain. After adjusting for all covariates, a unit increase in age decreased (Adjusted β-coef =-0.07 95% CI: -0.11, -0.04) the probability of having a high quality of life in the environmental domain. Females were more likely (Adjusted β-coef =0.99 95% CI: 0.54, 1.45) to have higher well-being in the environmental domain after adjusting for all other variables. (Table 5)

#### General Quality of life

A unit increase in age decreased (Crude β-coef = -0.02 95% CI: -0.02, -0.01) participants’ overall quality of life. Being a female increased (Crude β-coef =1.18 95% CI:0.68, 1.67) the probability of having a high quality of life. Similarly, being married increased (Crude β-coef =0.31 95% CI: 0.17, 0.46) the overall quality of life of participants compared to not being married. After adjusting for all other variables, a unit increase in age decreased (Adjusted β-coef =-0.04 95% CI: 0.05, -0.02) participants’ overall quality of life. Similarly, being ill health decreased (Adjusted β-coef =-2.00 95% CI: 8.13, 10.13) the probability of having a high quality of life among participants. (Table 5)

#### Pearson’s Correlation Matrix of the Variables of the Study

Table 6 shows the correlation matrix representing the linear relationship among the study variables. The study conducted a correlation analysis between the predictors of quality of life and the control variables understudy.

**Table 6:** Pearson’s Correlation Matrix of the Variables of the Study.

|  | 1 | 2 | 3 | 4 | 5 | 6 | 7 | 8 |  |
| --- | --- | --- | --- | --- | --- | --- | --- | --- | --- |
| Physical Domain | .35** | 1 |  |  |  |  |  |  |  |
| Psychological Domain | .32** | .39** | 1 |  |  |  |  |  |  |
| Social Domain | .36** | .42** | .38* | 1 |  |  |  |  |  |
| Environmental Domain | .46** | .43** | .51** | .43* | 1 |  |  |  |  |
| Age | .51** | .42** | .44** | .46** | .39* | 1 |  |  |  |
| Level of education | .56** | .32* | .32** | .47** | .52** | .48* | 1 |  |  |
| Gender | -.06 | .03 | -.05 | -.02 | .01 | .02 | .03 | 1 |  |
| Employment status | .02 | .10 | .11 | .05 | .00 | -.07 | .00 | .21 | 1 |

\*\**. Correlation is significant at the 0.01 level (2-tailed).*
\**. Correlation is significant at the 0.05 level (2-tailed).*

Regarding the correlation between the predicting and control variables, the results showed that age and level of education positively correlated with quality of life, which were conceptualized in the constructs such as physical, psychological, social, and environmental domains. That is, the age of the person and level of education have an impact on the quality of life of the patient. However, gender and years of working failed to correlate with quality of life.

## Discussion

The study assessed the health-related quality of life of individuals with type 2 diabetes in Ghana. The study compared WHQOL-BREF mean scores across four domains based on sex, age, education status, marital status, employment status, and health status. Participants under 50 exhibited higher mean scores in all four domains (physical, psychological, social, and environmental) of health and overall quality of health compared to participants aged 50 years and above.

This study discovered that males had higher mean scores for all four domains and the overall quality of life than females. For instance, males had a mean score of 22.52 and SD of 3.16 compared to the mean score and SD of 21.29±2.98 for females. A study in Western Ethiopia conducted among patients with diabetes to assess their HRQoL found a mean score of overall HRQoL 50.3+-18.1^13^. There was a high HRQoL (63.2+- 34.4) in the physical functioning domain and the lowest HRQoL (30.2+-22.9) in the general health domain^13^. The most affected domains of HRQoL with less than a mean score of 50 were general health, mental health, bodily pains, and vitality among patients with diabetes^13^. Another cross-sectional study on predictors of QoL and glycemic control among Saudi adults with diabetes found a higher QoL score of 74.1+11.6^20^. Also, a cross-sectional study on predictors of QoL and other patient-reported outcomes with people with type 2 diabetes indicates a good QoL of a score of 0.93, which approximates good^15^.

On the other hand, a cross-sectional survey study on the association between complications and HRQoL in individuals with diabetes in Singapore shows that diabetes patients with complications had lower HRQoL than those without complications^21^. Another study done in Ghana and Nigeria on predictors of QoL in patients with diabetes mellitus in two tertiary health institutions shows generally low (66.14+-9.99 and 68.78+-7.86) QoL in Ghana and Nigeria, respectively^16^. In a cross-sectional descriptive study done on profile and predictors of HRQoL among type 2 diabetes mellitus patients in Pakistan, poor (54.58+-20.28), HRQoL among type 2 diabetes patients was reported^17^. A related study on predictors of HRQoL in diabetic neuropathy in type 2 diabetes patients in Greece shows poor HRQoL^22^. Similarly, a cross-sectional survey of demographic and clinical predictors of HRQoL among people with type 2 diabetes in Thailand showed a poor HRQoL^23^. Health-related quality of life (HRQoL) is a multidimensional concept that reflects an individual’s overall well-being and perceived health status. It encompasses various physical, mental, and social aspects of a person’s life. It can have several implications across different domains: HRQoL assessments can inform healthcare providers about the impact of a disease or treatment on a patient’s overall well-being. This information can be crucial in making treatment decisions and assessing the effectiveness of interventions. Understanding a patient’s HRQoL allows healthcare professionals to provide more patient-centered care, tailoring treatment plans to individual needs and priorities.

HRQoL measures are increasingly used to evaluate the overall impact of treatments, interventions, or new medications on patients’ lives and traditional clinical outcomes^24^. HRQoL data can inform healthcare policy decisions by providing insights into the burden of diseases on individuals and society^25^. This can influence resource allocation and healthcare prioritization. Poor HRQoL can result in increased healthcare costs due to higher utilization of healthcare services. It can also lead to reduced work productivity and an economic burden on individuals and society. HRQoL measures can help identify psychological and social issues that affect an individual’s well-being, such as depression, anxiety, or social isolation^25,26^.

HRQoL assessments can guide public health initiatives by identifying populations with lower HRQoL and implementing targeted interventions.^25^. HRQoL results can be used to educate patients about the impact of their conditions on their daily lives, motivating them to make healthier choices and adhere to treatment plans^25,27^. Healthcare organizations can use HRQoL data to assess the quality of care they provide and identify areas for improvement, ultimately leading to better patient outcomes. HRQoL assessments can highlight disparities in health outcomes among different demographic groups, helping policymakers and healthcare providers address these inequities. HRQoL considerations are essential in long-term care planning, particularly for individuals with chronic conditions or disabilities, as they can impact decisions regarding assisted living, nursing homes, or home healthcare. HRQoL data can empower patient advocacy groups and support organizations to advocate for better care, research, and policies that improve the quality of life for individuals with specific health conditions^25^. Health-related quality-of-life assessments have wide-ranging implications across healthcare, research, policy, and individual well-being. They provide valuable insights into the holistic impact of health conditions and interventions, ultimately contributing to more informed and patient-centered healthcare decisions and policies.

### Factors influencing Health-Related Quality of Life (HRQoL) of T2DM Patients in Ghana

There was a statistically significant association between age, sex, educational status, marital status, employment status, and health status of study participants. A unit increase in age decreases (Crude β-coef = -0.25, 95% CI: 0.28, -0.21) the probability of having good physical health. Females were more likely (Crude β-coef = 1.07 95% CI: 0.54, 1.40) to have increased quality of life in the physical domain compared to their male counterparts. A unit increase in age decreases (Crude β-coef= -0.12 95% CI: -0.15 -0.09) psychological health of study participants. Similarly, being married increased (Crude β-coef = 1.69 95% CI: 1.18, 2.21) the quality of life in the psychological domain compared to not being married. Being employed increased (Crude β-coef =1.62 95% CI: 1.23, 2.01) the psychological health of participants compared to unemployed participants.

Being educated increased (Crude β-coef = 0.71 95% CI: 0.46, 0.96) the quality of life in the social domain compared to not being educated. Similarly, being married increased (Crude β-coef =0.74 95% CI: 0.49, 0.98) the quality of life of participants in the social domain compared to unmarried participants. Females were more likely to have increased (Crude β-coef = 1.18 95% CI: 0.68, 1.67) QoL in the domain of the environment compared to males. Being employed increased the likelihood (Crude β-coef =0.78 95% CI: 0.26, 1.31) of having a high quality of life in the environmental domain compared to unemployed participants. A unit increase in age decreased (Crude β-coef = -0.02 95% CI: -0.02, -0.01) the total QoL of participants. Being a female increased (Crude β-coef =1.18 95% CI:0.68, 1.67) the probability of having a high quality of life. Similarly, being married increased (Crude β-coef =0.31 95% CI: 0.17, 0.46) the overall quality of life of participants compared to not being married.

This is consistent with a cross-sectional survey study that shows diabetes patients with complications had lower HRQoL than those without complications^21^. Another study done in Ghana and Nigeria on predictors of QoL in patients with diabetes mellitus in two tertiary health institutions shows generally low (66.14+-9.99 and 68.78+-7.86) QoL in Ghana and Nigeria, respectively^16^. A cross-sectional descriptive study done on profile and predictors of HRQoL among type 2 diabetes mellitus patients in Pakistan reported poor (54.58+-20.28) HRQoL among type 2 diabetes patients^17^. A related study on predictors of HRQoL in diabetic neuropathy in type 2 diabetes patients in Greece shows poor HRQoL^22^. Similarly, a cross-sectional survey of demographic and clinical predictors of HRQoL among people with type 2 diabetes in Thailand showed a poor HRQoL^23^.

Some researchers explained that the main stumbling blocks to adherence revolve around the general issues that patients can control: the oversight on taking prescribed medicine, intentionally omitting doses, and failure to seek complete information from doctors and other healthcare workers on unclear treatment issues^28,29^. Besides, patients ‘emotional state also hinders full adherence to prescribed treatment regimens. A diabetic person who is depressed is more likely to abandon the treatment regimens recommended by the doctor. Therefore, constantly paying attention to them is essential to boosting adherence levels. Different studies conducted have also indicated that socio-demographics like gender, marital status, and age are great contributors to low adherence levels^28,30–32^.

Besides, these authors have constantly cited gender as a significant factor in the non-adherence and adherence levels among patients. A different study on adherence to diabetes treatment in Uganda reported that more females did not understand the prescribed regimen compared to their male counterparts^32^. However, other factors like marriage status, employment status, and age have not been adequately related to low adherence levels to diabetes treatment regimens^32,33^. In a few cases, literacy levels have also been associated with better adherence tendencies.

Regardless, there is a contradiction to the findings. A study in Western Ethiopia conducted among patients with diabetes to assess their HRQoL found a mean score of overall HRQoL 50.3+-18.1^13^. There was a high HRQoL (63.2+- 34.4) in the physical functioning domain and the lowest HRQoL (30.2+-22.9) in the general health domain^13^. The most affected domains of HRQoL with less than a mean score of 50 are public health, mental health, bodily pains, and vitality among patients with diabetes^13^. Another cross-sectional study on predictors of QoL and glycemic control among Saudi adults with diabetes found a higher QoL score of 74.1+11.6^14^. Also, a cross-sectional study on predictors of QoL and other patient-reported outcomes with people with type 2 diabetes indicates a good QoL of a score of 0.93, which approximates good^15^.

For the predictors of the study, the findings of this study are in support of an earlier cross-sectional study carried out on predictors of HRQoL among patients with diabetes on follow-up in Western Ethiopia, indicating that a unit increase in age is likely to decrease HRQoL of the patient with diabetes by 0.25^13^. The male diabetes patients had about five times better HRQoL than females^13^. Diabetes patients who were married had five times better HRQoL compared to unmarried patients, and those who were unable to read and write had about nine times lower HRQoL, 3.6 units compared with those who could read and write^13^. History of smoking causes nine units lower HRQoL compared to their counterparts without a history of smoking, absence of comorbidity, and chronic complications related to diabetes cause an increase in HRQoL, and any unit increase in body mass index (BMI) is likely to decrease HRQoL by 3.56.^13^

Another cross-sectional study done in Ghana and Nigeria on predictors of QoL in patients with diabetes mellitus in two tertiary health institutions shows that the main predictors of QoL for diabetes are medication adherence, employment status, and diabetes empowerment^16^. A cross-sectional descriptive study done on the profile and predictors of HRQOL among type 2 diabetes mellitus patients in Pakistan reported that higher medication adherence was associated with improvement in HRQOL for type 2 diabetes patients^17^. Another cross-sectional observational study on predictors of HRQOL among patients with type 2 diabetes who are using insulin shows a significant association between the age, treatment satisfaction, medication adherence, BMI, and HbA1c of an individual and predictors of HRQOL for type 2 diabetes patients using insulin^18^.

In a cohort study on temporal predictors of HRQoLin in elderly people with diabetes in Germany, the study found that diabetes-related complications, higher BMI, and smoking are strong predictors of HRQoL in elderly patients with diabetes^34^. Similarly, a cross-sectional survey of demographic and clinical predictors of HRQoL among people with type 2 diabetes in Thailand found that diabetic foot ulcers and smoking status were significant predictors of low HRQoL among type 2 diabetes patients^23^. Another, a cross-sectional study on predictors of QoL life and glycemic control among Saudi adults with diabetes found men had higher QoL than women, patients who are educated have better QoL than those without education, and there is a high prevalence of poor glycemic control among men than women^14^.

This finding aligns with the understanding that functional literacy is crucial regarding health information. In Sub-Saharan Africa (SSA), there’s limited recognition of the burden of chronic non-communicable diseases like diabetes^35^. Health literacy is a person’s capacity to seek, comprehend, and apply health information to improve health outcomes^36^. Furthermore, the study indicated that individuals face various challenges in accessing healthcare services, some of which are not related to health insurance but instead to health literacy. They recommend enhancing accessible health information online^36^.

Predictors of quality of life among individuals with diabetes are factors or variables that can influence the overall well-being and perceived health status of people living with this chronic condition. Understanding these predictors can help healthcare providers, researchers, and policymakers develop targeted interventions and support strategies to address specific challenges faced by individuals with diabetes to improve their quality of life.

### Limitation

This study is limited to only three health facilities in three selected regions out of the 16 regions in Ghana. This means that the study findings might not apply to other regions in Ghana.

Secondly, the study adopted a cross-sectional design, restricting our ability to establish causation or track changes over time.

## Conclusions and Recommendations

This study provided valuable insights on predictors of health-related QoL among type 2 diabetic patients in Ghana, which would guide policymakers, healthcare providers, and stakeholders in improving diabetes management and ultimately enhancing the quality of life for individuals living with diabetes in Ghana. Males have a better quality of life than females, and those below 50 also have a better quality of life than those above 50.

Evidence-based interventions should be implemented to address the factors influencing the QoL of diabetic patients in Ghana. The various health facilities must develop diabetes education programs to empower patients with the knowledge and skills to manage their condition effectively and encourage self-care and self-monitoring to improve their quality of life.

## Data Availability

All data produced in the present work are contained in the manuscript

## Acknowledgements

The authors acknowledge all participants for their time and cooperation during the study.

## Notes

### Competing Interest Statement

The authors have declared no competing interest.

### Funding Statement

This study did not receive any funding

### Author Declarations

Ethical clearance was obtained from the Ghana Health Service Ethics Review Committee and the University of Port Harcourt Research Ethics Review Board

## Reference

1. WHO. The top 10 causes of death - Factsheet. WHO reports. 2020;(December 2020).

2. CDC. Diabetes Quick Facts | Basics | Diabetes. Centers for Disease Control and Prevention.

3. Khan MAB, Hashim MJ, King JK, Govender RD, Mustafa H, Kaabi J Al. Epidemiology of Type 2 diabetes - Global burden of disease and forecasted trends. J Epidemiol Glob Health. 2020;10(1). doi:10.2991/JEGH.K.191028.001

4. Dorvlo GGK, Kumah A, Ofosu SK, et al. Factors Associated with Antidiabetic Medications and Dietary Recommendation Adherence Among Patients with Type 2 Diabetes. Global Journal on Quality and Safety in Healthcare. Published online June 17, 2024. doi:10.36401/jqsh-23-52

5. Adeloye D, Ige JO, Aderemi A V., et al. Estimating the prevalence, hospitalisation and mortality from type 2 diabetes mellitus in Nigeria: A systematic review and meta-analysis. BMJ Open. 2017;7(5). doi:10.1136/bmjopen-2016-015424

6. Bawah AT, Ngambire LT, Abaka-Yawson A, Anomah A, Kinanyok S, Tornyi H. A community based prevalence of type 2 diabetes mellitus in the Ho municipality of Ghana. Journal of Public Health (Germany). 2021;29(2). doi:10.1007/s10389-019-01144-7

7. Gudjinu HY, Sarfo B. Risk factors for type 2 diabetes mellitus among out-patients in Ho, the Volta regional capital of Ghana: A case-control study. BMC Res Notes. 2017;10(1). doi:10.1186/s13104-017-2648-z

8. Addo J, Agyemang C, de-Graft Aikins A, et al. Association between socioeconomic position and the prevalence of type 2 diabetes in Ghanaians in different geographic locations: The RODAM study. J Epidemiol Community Health (1978). 2017;71(7). doi:10.1136/jech-2016-208322

9. Sarfo-Kantanka O, Sarfo FS, Ansah EO, Eghan B, Ayisi-Boateng NK, Acheamfour-Akowuah E. Secular trends in admissions and mortality rates from diabetes mellitus in the Central Belt of Ghana: a 31-year review. Pan African Medical Journal Conference Proceedings. 2018;1. doi:10.11604/pamj.cp.2017.2.38.72

10. Annani-Akollor ME, Addai-Mensah O, Fondjo LA, et al. Predominant complications of type 2 diabetes in kumasi: A 4-year retrospective cross-sectional study at a teaching hospital in ghana. Medicina (Lithuania). 2019;55(5). doi:10.3390/medicina55050125

11. Hanefeld J, Powell-Jackson T, Balabanova D. Understanding and measuring quality of care: dealing with complexity. Bull World Health Organ. 2017;95(5). doi:10.2471/blt.16.179309

12. WHO. WHO | WHOQOL: Measuring Quality of Life. Health statistics and information systems (WHO). Published online 2020.

13. Feyisa BR, Yilma MT, Tolessa BE. Predictors of health-related quality of life among patients with diabetes on follow-up at Nekemte specialised Hospital, Western Ethiopia: A cross-sectional study. BMJ Open. 2020;10(7). doi:10.1136/bmjopen-2019-036106

14. Huda MD, Rahman M, Rahman MM, Islam MJ, Haque SE, Mostofa MG. Readiness of health facilities and determinants to manage diabetes mellitus: Evidence from the nationwide Service Provision Assessment survey of Afghanistan, Bangladesh and Nepal. BMJ Open. 2021;11(12). doi:10.1136/bmjopen-2021-054031

15. Bradley C, Eschwège E, De Pablos-Velasco P, et al. Predictors of quality of life and other patient-Reported outcomes in the PANORAMA multinational study of people with type 2 diabetes. Diabetes Care. 2018;41(2). doi:10.2337/dc16-2655

16. Ababio GK, Bosomprah S, Olumide A, et al. Predictors of quality of life in patients with diabetes mellitus in two tertiary health institutions in Ghana and Nigeria. Niger Postgrad Med J. 2017;24(1). doi:10.4103/npmj.npmj_3_17

17. Iqbal Q, ul Haq N, Bashir S, Bashaar M. Profile and predictors of health related quality of life among type II diabetes mellitus patients in Quetta city, Pakistan. Health Qual Life Outcomes. 2017;15(1). doi:10.1186/s12955-017-0717-6

18. Gillani SW, Ansari IA, Zaghloul HA, et al. Predictors of Health-Related Quality of Life Among Patients with Type II Diabetes Mellitus Who Are Insulin Users: A Multidimensional Model. Curr Ther Res Clin Exp. 2019;90. doi:10.1016/j.curtheres.2019.04.001

19. WHO. WHOQOL: measuring quality of life. Psychol Med. 1998;28(3).

20. Nazir SUR, Hassali MA, Saleem F, Bashir S, Hashmi F, Aljadhey H. A cross-sectional assessment of health-related quality of life among type 2 diabetic patients in Pakistan. J Pharm Bioallied Sci. 2016;8(1). doi:10.4103/0975-7406.171734

21. Venkataraman K, Wee HL, Leow MKS, et al. Associations between complications and health-related quality of life in individuals with diabetes. Clin Endocrinol (Oxf). 2013;78(6). doi:10.1111/j.1365-2265.2012.04480.x

22. Georgios Lyrakos N, Hatziagelaki E, Damigos D, Athanasia Papazafiropoulou K, Bousboulas S, Batistaki C. Predictors of health-related quality of life in diabetic neuropathy type II diabetic patients in Greece. Health Science Journal. 2013;7(3).

23. Khunkaew S, Fernandez R, Sim J. Demographic and clinical predictors of health-related quality of life among people with type 2 diabetes mellitus living in northern Thailand: A cross-sectional study. Health Qual Life Outcomes. 2019;17(1). doi:10.1186/s12955-019-1246-2

24. Yin S, Njai R, Barker L, Siegel PZ, Liao Y. Summarizing health-related quality of life (HRQOL): Development and testing of a one-factor model. Popul Health Metr. 2016;14(1). doi:10.1186/s12963-016-0091-3

25. Kaplan RM, Hays RD. Health-Related Quality of Life Measurement in Public Health. Annu Rev Public Health. 2022;43. doi:10.1146/annurev-publhealth-052120-012811

26. Yildirim G, Rashidi M, Karaman F, et al. The relationship between diabetes burden and health-related quality of life in elderly people with diabetes. Prim Care Diabetes. 2023;17(6). doi:10.1016/j.pcd.2023.08.007

27. Oluchi SE, Manaf RA, Ismail S, Kadir Shahar H, Mahmud A, Udeani TK. Health related quality of life measurements for diabetes: A systematic review. Int J Environ Res Public Health. 2021;18(17). doi:10.3390/ijerph18179245

28. Wexler DJ, Grant RW, Wittenberg E, et al. Correlates of health-related quality of life in type 2 diabetes. Diabetologia. 2006;49(7). doi:10.1007/s00125-006-0249-9

29. Solli O, Stavem K, Kristiansen IS. Health-related quality of life in diabetes: The associations of complications with EQ-5D scores. Health Qual Life Outcomes. 2010;8. doi:10.1186/1477-7525-8-18

30. Adisa R, Alutundu MB, Fakeye TO. Factors contributing to nonadherence to oral hypoglycemic medications among ambulatory type 2 diabetes patients in Southwestern Nigeria. Pharm Pract (Granada). 2009;7(3). doi:10.4321/S1886-36552009000300006

31. Kirkman MS, Briscoe VJ, Clark N, et al. Diabetes in older adults. Diabetes Care. 2012;35(12). doi:10.2337/dc12-1801

32. Kalyango JN, Hall M, Karamagi C. Home medication management practices and associated factors among patients with selected chronic diseases in a community pharmacy in Uganda. BMC Health Serv Res. 2012;12(1). doi:10.1186/1472-6963-12-323

33. Gálvez Galán I, Cáceres León MC, Guerrero-Martín J, López Jurado CF, Durán-Gómez N. Health-related quality of life in diabetes mellitus patients in primary health care. Enferm Clin. 2021;31(5). doi:10.1016/j.enfcli.2021.03.001

34. Maatouk I, Wild B, Wesche D, et al. Temporal predictors of health-related quality of life in elderly people with diabetes: Results of a German cohort study. PLoS One. 2012;7(1). doi:10.1371/journal.pone.0031088

35. Nuche-Berenguer B, Kupfer LE. Erratum: Readiness of Sub-Saharan Africa healthcare systems for the new pandemic, diabetes: A systematic review (Journal of Diabetes Research (2018) 2018 (9262395) DOI: 10.1155/2018/9262395). J Diabetes Res. 2018;2018. doi:10.1155/2018/3419290

36. Batterham RW, Buchbinder R, Beauchamp A, Dodson S, Elsworth GR, Osborne RH. The OPtimising HEalth LIterAcy (Ophelia) process: Study protocol for using health literacy profiling and community engagement to create and implement health reform. BMC Public Health. 2014;14(1). doi:10.1186/1471-2458-14-694

